# Effectiveness of Relaxation Interventions on Anxiety, Depression, and Quality of Life in Women with Infertility Undergoing Assisted Reproductive Technology: A Meta-Analysis of Controlled Trials

**DOI:** 10.64898/2026.02.26.26347155

**Authors:** Seo A Park, Hye young Kim

**Author notes:** **Corresponding author**: Hye young Kim, PhD, RN.

## Abstract

This systematic review and meta-analysis aimed to evaluate the effectiveness of relaxation interventions on anxiety, depression, stress, and quality of life in women with infertility. A comprehensive search of PubMed, OVID MEDLINE, CINAHL®, Google Scholar, and Korean databases was conducted for articles published through March 2025. Keywords included combinations of terms related to infertility, ART, and nursing or psychotherapeutic interventions. The search identified 759 records, of which 13 met the eligibility criteria. Methodological quality was assessed using the Cochrane Risk of Bias tool, and data analysis was performed using R software (version 4.3.2). The meta-analysis included 10 randomized controlled trials (RCTs) and three non-randomized controlled trials (NRCTs), comprising 1,215 women undergoing ART. Intervention groups received relaxation programs, while comparison groups received usual care or no intervention. Relaxation interventions were associated with significant reductions in anxiety (Hedges’ *g* = −0.69) and depression (Hedges’ *g* = −0.38), and significant improvements in quality of life (Hedges’ *g* = 0.25). No statistically significant effect was observed for stress (Hedges’ *g* = −0.01; 95% CI: −0.49 to 0.47). Heterogeneity and risk of publication bias were determined to be low. Overall, relaxation programs demonstrated beneficial effects on anxiety, depression, and quality of life, but not on stress levels. Relaxation interventions appear to support the psychological well-being of women undergoing ART, with particular benefit for women with a history of repeated treatment failure. Individualized, woman-centered approaches may be more responsive to the needs of this population than universal or group-based models of care.

## 1. Introduction

Amidst declining global fertility rates, South Korea reported a total fertility rate (TFR) of 0.72 in 2023, representing the lowest rate among Organization for Economic Co-operation and Development (OECD) member countries and globally [1–2]. This persistently low fertility rate constitutes a critical national social issue, necessitating effective policy and health interventions. Infertility directly impacts pregnancy and childbirth and is a primary contributor to declining birth rates [3–4]. Clinically, infertility is defined as the failure to establish a pregnancy after 1 year of regular, unprotected sexual intercourse in women and men aged 15 to 49 years [5]. Globally, an estimated 60 to 80 million couples experience difficulty conceiving [4,6]. The etiology of infertility is attributed to female factors in approximately 40%–50% of cases, male factors in 35%–45%, combined factors in approximately 10%, and unexplained causes in 5%–10% of cases [7–9]. Infertility represents a family developmental crisis affecting both partners; however, women report significantly higher levels of psychological stress than men [10]. Consequently, infertility treatment requires not only biomedical approaches but also multidimensional medical and psychosocial support that addresses the physical, psychological, social, and environmental challenges faced by women with infertility.

Advances in medical technology have facilitated the widespread use of assisted reproductive technology (ART), resulting in increased rates of successful pregnancies [11]. Despite these advances, women with infertility undergoing ART often experience exacerbated physical, psychological, and economic burdens throughout the treatment process [10]. During treatment, women frequently report adverse emotional responses, including anxiety, depression, anger, isolation, sexual dysfunction, reduced self-esteem, and heightened stress regarding uncertain pregnancy outcomes, alongside substantial financial strain associated with treatment costs [10,12–18]. Furthermore, repeated diagnostic examinations and treatment procedures aimed at identifying reproductive health pathologies further intensify physical and emotional stress, contributing to depressive symptoms [14,17,19–20]. These cumulative adverse experiences negatively affect ART outcomes and pregnancy success rates, ultimately diminishing the quality of life of women with infertility [12,15,21–22].

Therefore, interventions that effectively attenuate anxiety, depression, and stress while enhancing quality of life in women undergoing ART are of considerable clinical importance. Mind–body relaxation programs are increasingly utilized as non-pharmacological interventions to address the physical, emotional, psychological, and social difficulties experienced by women receiving ART [23]. Studies indicate that these programs alleviate adverse emotions and emotional distress and may improve the likelihood of pregnancy [24]. Mind–body relaxation interventions encompass a range of techniques, including breathing exercises, guided imagery, meditation, progressive muscle relaxation, and music therapy [25], and previous research has demonstrated their effectiveness in reducing stress, anxiety, and depressive symptoms among women with infertility [10,26–27]. Physiologically, relaxation responses are associated with increased parasympathetic nervous system activity and decreased sympathetic activation, leading to reductions in blood pressure and heart rate [25], pain relief [28], and decreased anxiety and stress levels [25].

Although recent studies have examined the effectiveness of mind–body relaxation interventions, evidence specific to women with infertility undergoing ART remains limited. In particular, few studies have systematically compared individual effect sizes, intervention strategies, and methodological differences across trials, limiting the translation of these interventions into routine clinical practice for ART populations. Accordingly, this study systematically reviewed domestic and international literature on mind–body relaxation intervention programs aimed at reducing anxiety, depression, and stress and improving quality of life in women with infertility undergoing ART. By estimating pooled effect sizes, this study aims to provide practical, evidence-based recommendations for future clinical practice and research.

## 2. Participants, Ethics and Methods

### 2.1. Literature search

This systematic review adhered to the *Systematic Literature Review Manual* of the National Evidence-based Healthcare Collaborating Agency (NECA) [29] and the Preferred Reporting Items for Systematic Reviews and Meta-Analyses (PRISMA) guidelines [30]. The study selection process was conducted in accordance with the PRISMA framework (Figure 1).

**Figure 1.**
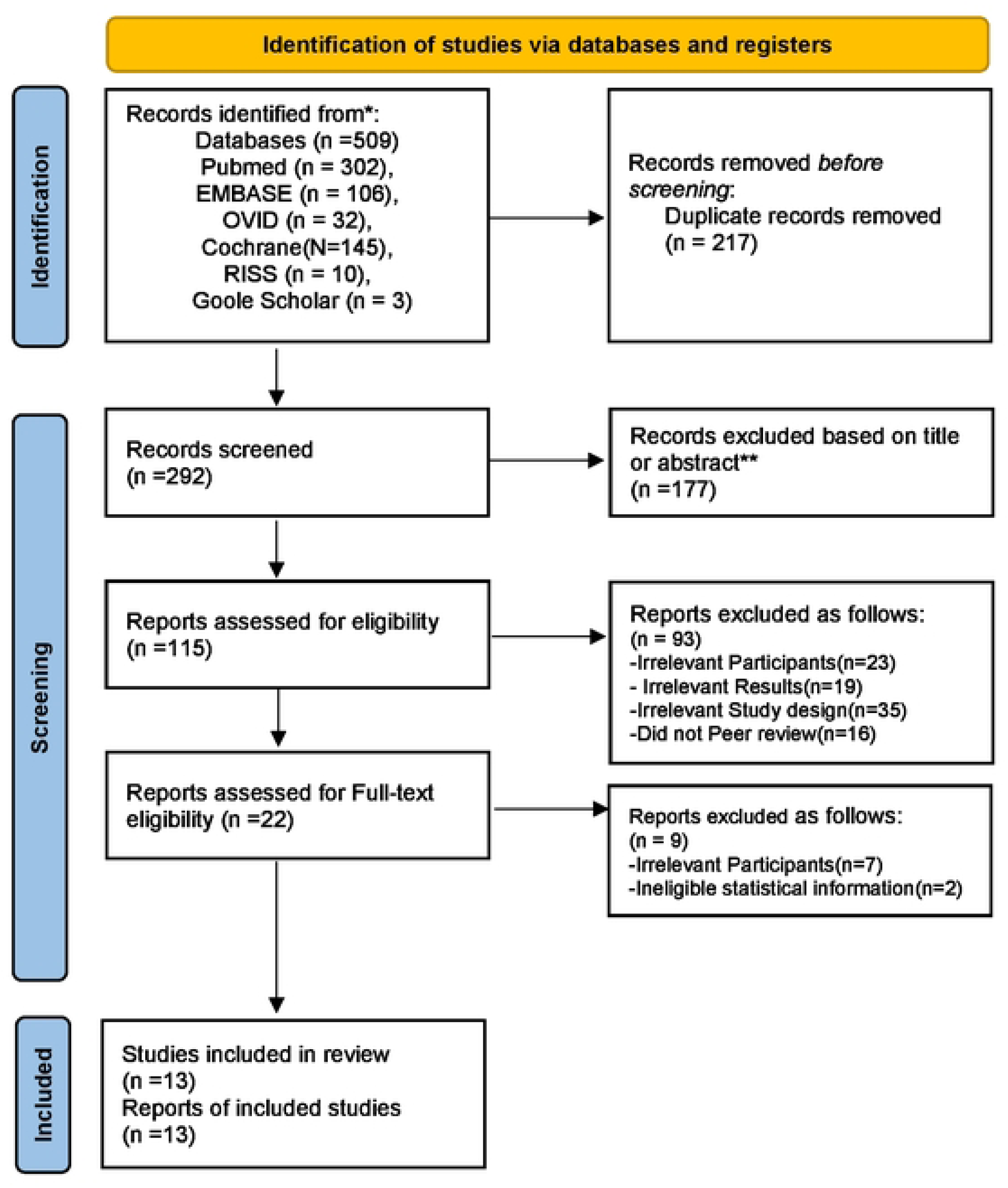
PRISMA flow chart of selection of studies.

### 2.2 Eligibility criteria

Selection criteria were defined using the PICO (Population, Intervention, Comparison, Outcome) framework. The population (P) comprised women diagnosed with infertility undergoing assisted reproductive technology (ART). The intervention (I) involved relaxation programs. The comparison (C) group received usual care, routine nursing, education, or conservative treatment. The outcomes (O) of interest were anxiety, depression, stress, and quality of life. Eligible studies included randomized controlled trials (RCTs) and non-randomized controlled trials (NRCTs) that reported the statistical data required to calculate effect sizes for both experimental and control groups, including means and standard deviations.

Inclusion criteria were: (1) studies involving women undergoing ART following an infertility diagnosis; (2) studies in which a relaxation program was applied to the experimental group; (3) studies in which the control group received usual care or conservative treatment; and (4) studies reporting at least one outcome related to anxiety, depression, stress, or quality of life. Exclusion criteria were: (1) non-randomized controlled trials (unless specified as NRCTs meeting quality criteria); (2) grey literature; and (3) non-peer-reviewed studies.

### 2.3. Search strategy and study selection

To identify studies that applied various relaxation intervention programs to infertile women undergoing assisted reproductive technology (ART), both international and domestic academic databases were searched using internet-based platforms. No restrictions were placed on the publication period, and the literature search was conducted up to March 2025.

International databases included Ovid MEDLINE and PubMed. Medical Subject Headings (MeSH) terms, synonyms, and related terms representing infertility, relaxation programs, and nursing interventions were identified. The search strategy combined the following terms: (((Infertility* OR Female) AND (Reproductive Techniques, Assisted OR Ovulation Induction OR Fertilization in Vitro OR Intrauterine Insemination OR Intracytoplasmic Sperm Injection)) AND ((Nursing AND (Psychotherapeutic OR Relaxation OR Cognitive Behavioral OR Mindfulness OR Intervention* OR Program*)) AND (Infertility Treatment* OR Nursing Intervention* OR Evaluation* OR Program*))). Domestic databases, including the Research Information Service System (RISS) and Google Scholar, were searched for published academic articles. Additional studies were identified through manual searching. Because domestic databases do not support MeSH-based searching, keywords were derived primarily from the terms “program” and “intervention” to enhance specificity. Keywords such as “assisted reproductive technology,” “infertility,” “women with infertility,” “relaxation therapy,” “nursing education,” “nursing intervention,” “psychological relaxation education program,” “psychological relaxation intervention,” and “psychotherapy” were used. All retrieved records were managed using reference management software. The electronic search yielded 509 records (496 international and 13 domestic). Following the removal of 217 duplicates, 292 records remained. Screening of titles and abstracts resulted in the exclusion of 177 records. Subsequently, 93 full-text articles were excluded for the following reasons: the population did not comprise women with infertility undergoing ART (*n* = 23); outcome variables did not include anxiety, depression, stress, or quality of life (*n* = 19); inappropriate study design (*n* = 35); or non-peer-reviewed status (*n* = 16). Full-text review was conducted for the remaining 22 articles. Of these, nine were excluded due to missing statistical data required for effect size calculation (*n* = 2) or other eligibility issues (*n* = 7). Ultimately, 13 studies met the inclusion criteria and were included in the final meta-analysis. Study selection was conducted independently by two researchers, with discrepancies resolved through consensus.

### 2.4. Quality assessment

Methodological quality was assessed using standardized risk-of-bias tools appropriate to the study design. RCTs were evaluated using the Cochrane Collaboration’s Risk of Bias (RoB) tool [31–32], while NRCTs were assessed using the Risk of Bias Assessment Tool for Non-randomized Studies (ROBANS) [33–34]. The RoB tool assessed potential bias across domains, including random sequence generation, allocation concealment, blinding of participants and personnel, blinding of outcome assessment, completeness of outcome data, selective reporting, and other sources of bias. The ROBANS tool evaluated group comparability, participant selection, confounding variable control, exposure measurement, blinding of outcome assessors, outcome assessment, incomplete outcome data, and selective reporting. Each domain was rated as having a low, high, or unclear risk of bias. Risk-of-bias assessment was conducted independently by two reviewers, with disagreements resolved through discussion.

### 2.5. Data analysis

Data extracted included author, publication year, country, research topic, study design, sample size, treatment modality, intervention characteristics (type, session frequency, duration, and session length), and outcome variables (anxiety, stress, depression, and quality of life). Pooled effect sizes were analyzed using R software (version 3.5.2; Meta-analysis with R). For continuous outcome variables, mean differences between experimental and control groups were estimated using the corrected standardized mean difference (Hedges’ *g*) with 95% confidence intervals (CIs) [35–37]. A random-effects model was applied to calculate pooled effect sizes to account for diversity in study design, sample composition, and intervention type. Statistical heterogeneity was assessed using the *Q* statistic, its corresponding *p* value, and the *I*² statistic. Heterogeneity was interpreted as low when *I*² ≤ 25%, moderate when *I*² > 25% and ≤ 75%, and high when *I*² > 75% [38]. Subgroup analyses using meta-analysis of variance (ANOVA) were performed based on moderator variables to explore sources of heterogeneity. Publication bias was examined using funnel plots.

## 3. Findings

### 3.1. Characteristics of included studies and participants

This review examined the characteristics of 13 studies evaluating relaxation interventions for women diagnosed with infertility undergoing ART (Table 1). A total of 1,215 participants were included. The majority of studies were conducted in Asia (*n* = 11; 84.6%), including four in South Korea [19, 20, 39, 40] (30.8%), two in Iran [17, 21] (15.4%), two in Turkey [18, 41] (15.4%), and one each in China [42] (7.7%), Japan [43] (7.7%), Hong Kong [44] (7.7%), and Taiwan [45] (7.7%). One study was conducted in Europe (Spain) [22] (7.7%). Regarding study design, 10 studies [17–18, 21–22, 39, 41–45] (76.9%) employed randomized controlled trial (RCT) designs, while three studies [19, 20, 40] (23.1%) were non-randomized controlled trials (NRCTs). The predominance of RCTs suggests that the overall body of evidence provides a relatively high level of methodological rigor.

Sample sizes varied considerably across the included studies. The smallest study enrolled 24 participants (12 in the intervention group and 12 in the control group) [39], whereas the largest study included 186 participants (89 in the intervention group and 97 in the control group) [41]. Regarding treatment modalities, all included studies involved *in vitro* fertilization (IVF) as the ART method. The most frequently applied interventions were mindfulness-based programs (*n* = 4) [19, 21, 40, 42]. Cognitive behavioral therapy (CBT)-based programs were utilized in three studies [22, 39, 45]. Additionally, relaxation-based interventions were applied in four studies [18, 22, 40, 45]. Specifically, one study combined progressive muscle relaxation with laughter therapy [18], one applied guided imagery [40], one implemented music therapy [41], and one utilized self-hypnosis combined with muscle relaxation [45]. Two studies employed integrative interventions incorporating physical, psychosocial, and spiritual components [17, 44]. One study applied a self-managed stress management program using a guidebook and diary [43]. Regarding delivery methods, individual-based interventions were conducted in nine studies [18–20, 22, 39–41, 43, 45], while group-based interventions were used in four studies [17, 21, 42, 44]. Analysis of intervention session duration indicated that two studies involved sessions lasting less than 30 min [40, 41], four studies reported durations of 40 to 120 min [17, 18, 22, 43], and six studies involved sessions lasting 120 to 240 min [19–21, 39, 42, 44].

Anxiety was the most frequently assessed psychological outcome, evaluated in 10 studies [17–19, 22, 39, 40, 41, 43–45]. Depression was assessed in seven studies [17–18, 20, 22, 39, 43, 45], and quality of life was evaluated in six studies [17, 21–22, 39, 42, 43]. Regarding measurement instruments, anxiety was most commonly assessed using the State–Trait Anxiety Inventory (STAI) (*n* = 5) [18–19, 22, 39, 41]. Additional instruments included the Visual Analogue Scale for Anxiety (VAS-A) in two studies [19, 40], the State Anxiety Inventory (SAI) in two studies [44, 45], and the Hospital Anxiety and Depression Scale (HADS) and Depression Anxiety Stress Scale–21 (DASS-21) in one study each [43]. Depression was measured primarily using the Beck Depression Inventory (BDI) (*n* = 3) [18, 22, 39], while the Centre for Epidemiologic Studies Depression Scale (CES-D) [20], HADS [43], and Self-Rating Depression Scale (SDS) [45] were each used in one study. Quality of life was assessed using the Fertility Quality of Life (FertiQoL) scale in four studies [22, 39, 42, 43], with the SF-36 [21] and SF-12 [17] used in one study each.

### 3.2. Quality assessment of included studies

Of the 13 studies included in the final analysis, 10 were RCTs. In all included studies, research objectives were clearly stated, homogeneity between the experimental and control groups was assessed, and measurement instruments with established reliability and validity were used. Appropriate statistical methods were applied in all analyses, and no participant attrition was reported. Among the 10 RCTs, one study [39] was rated as having an ‘unclear’ risk of bias due to inadequate reporting of allocation concealment. Three studies were identified as NRCTs [19, 20, 40]. In all three studies, research objectives were clearly defined, comparability between the experimental and control groups was ensured, and validated measurement tools were used. The attrition rate across these NRCTs was less than 20%. Furthermore, the risk of bias was assessed as low for all other domains, including performance bias and detection bias related to inappropriate intervention implementation or exposure measurement. Based on the comprehensive quality assessment, the overall risk of bias for the 13 selected studies was judged to be low (Figure 2).

**Figure 2.**
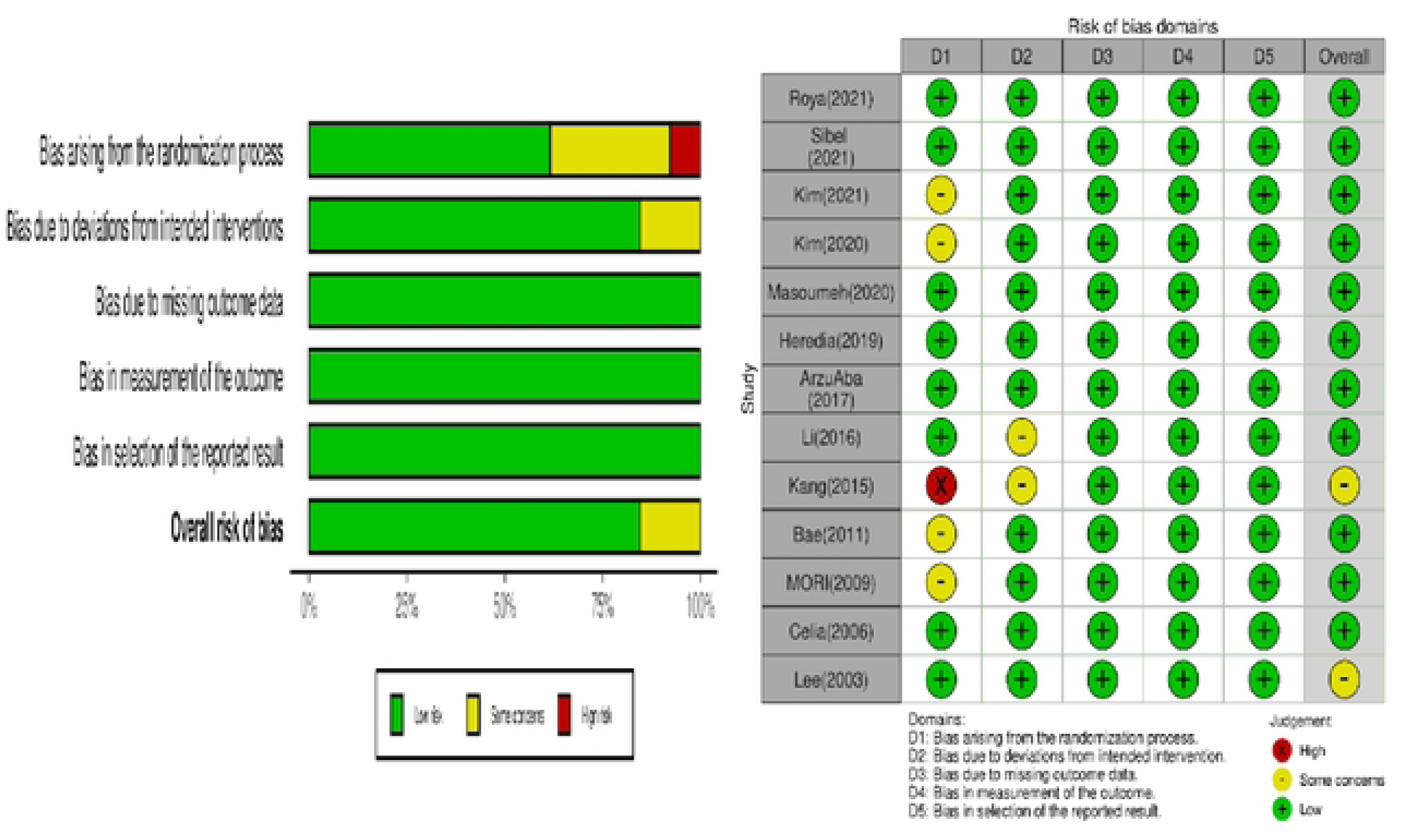
Risk of bias graph.

### 3.3. Effectiveness of Relaxation Programs

This meta-analysis included 13 studies applying mind–body relaxation programs to women with infertility undergoing ART. Outcome variables were categorized into anxiety, depression, stress, and quality of life. For each outcome, standardized mean differences (Hedges’ *g*) were calculated using the means, standard deviations, and sample sizes of the experimental and control groups. Pooled effect sizes were presented using forest plots (Figure 3).

**Figure 3.**
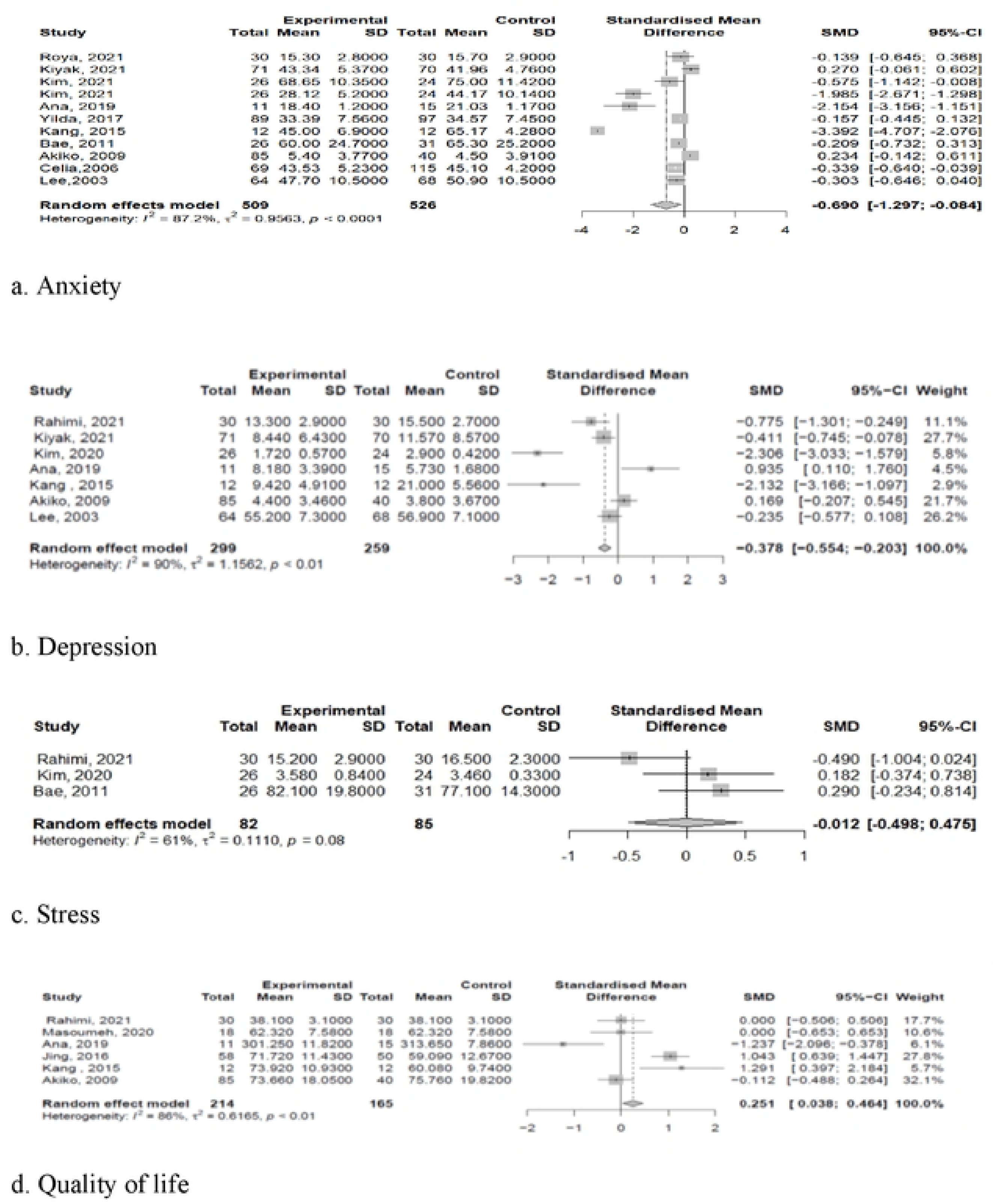
Forest plots of the effects of relaxation-intervention program.

#### 3.3.1. Anxiety

Pooled analysis demonstrated that relaxation programs had a statistically significant effect on anxiety, with a moderate-to-large effect size (Hedges’ *g* = −0.69; 95% CI: −1.30 to −0.08; *p* = .026). The heterogeneity assessment indicated a high degree of heterogeneity among studies (*I*² = 87.2%; *p* < .001).

#### 3.3.2. Depression

Relaxation programs also demonstrated a statistically significant reduction in depression, with a pooled effect size of Hedges’ *g* = −0.37 (95% CI: −0.55 to −0.20; *p* < .001). Heterogeneity among studies was high (*I*² = 89.8%; *p* < .001).

#### 3.3.3. Stress

In contrast, the pooled effect size for stress was statistically non-significant (Hedges’ *g* = −0.01; 95% CI: −0.49 to 0.47; *p* = .64). Heterogeneity analysis indicated a moderate to high level of heterogeneity (*I*² = 61%; *p* = .08).

#### 3.3.4. Quality of life

For quality of life, relaxation programs showed a small but statistically significant positive effect, with a pooled effect size of Hedges’ *g* = 0.25 (95% CI: 0.03 to 0.46; *p* = .02). Heterogeneity among studies was high (*I*² = 86.3%; *p* < .001).

### 3.4. Moderator analysis

Of the 10 studies assessing anxiety [17–19, 22, 39, 40, 41, 43–45], five evaluated anxiety using multiple indicators, such as state–trait anxiety, HADS, DASS-21, C-SAI, and visual analogue scales [18, 19, 41, 44, 45]. Overall, mind–body relaxation programs were associated with a statistically significant reduction in anxiety in the experimental group compared with the control group (Hedges’ *g* = −0.69; 95% CI: −1.30 to −0.08; *p* = .026). However, substantial heterogeneity was observed among the included studies (*Q* = 78.38; *p* < .001; *I*² = 87.2%), indicating considerable variability in effect sizes. This finding necessitated exploratory analysis to identify potential sources of heterogeneity and to determine which study characteristics contributed to differences in intervention effects. Accordingly, subgroup analyses were conducted using study design, delivery mode, intervention duration, number of sessions, and treatment status as moderator variables (Figure 4).

**Figure 4.**
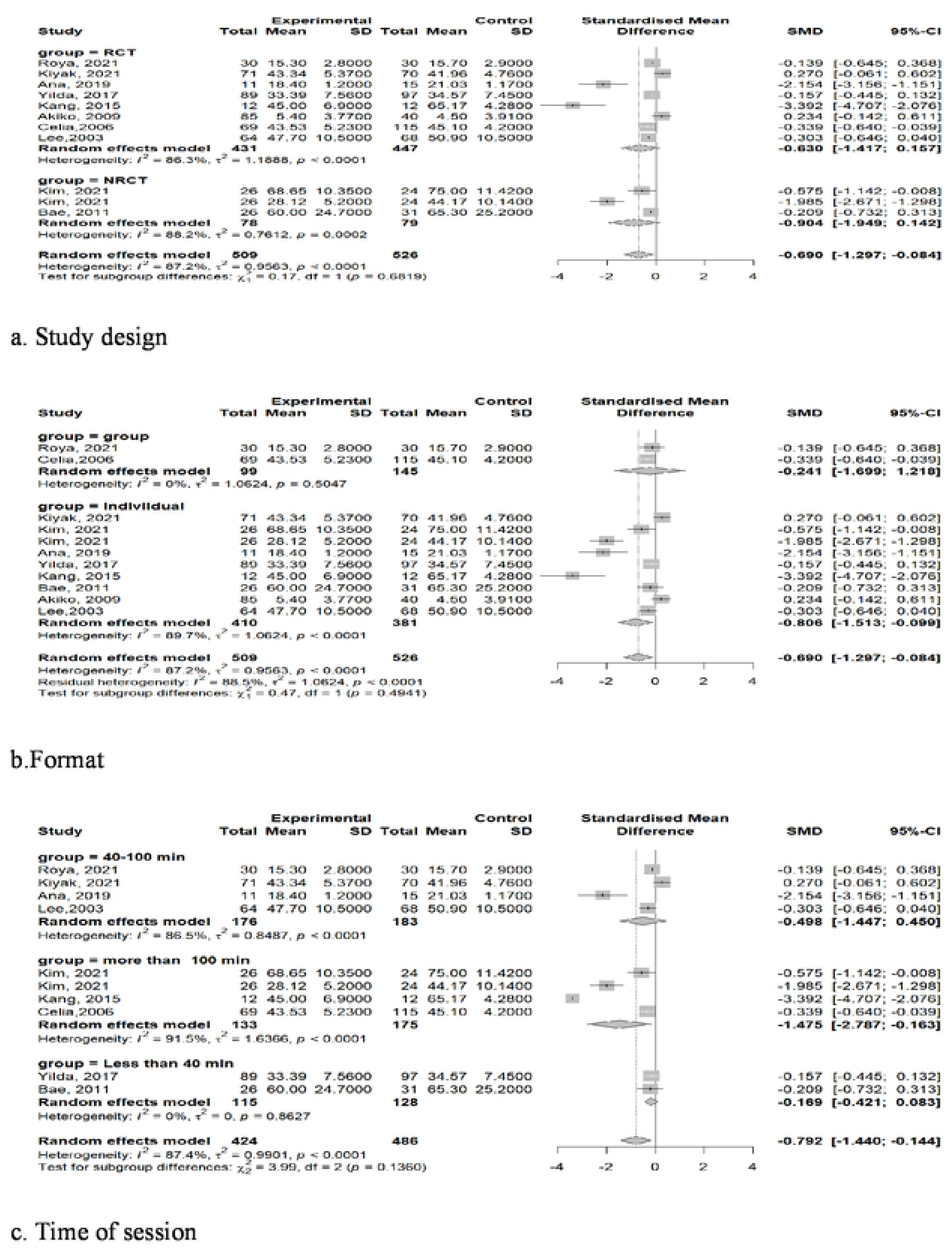
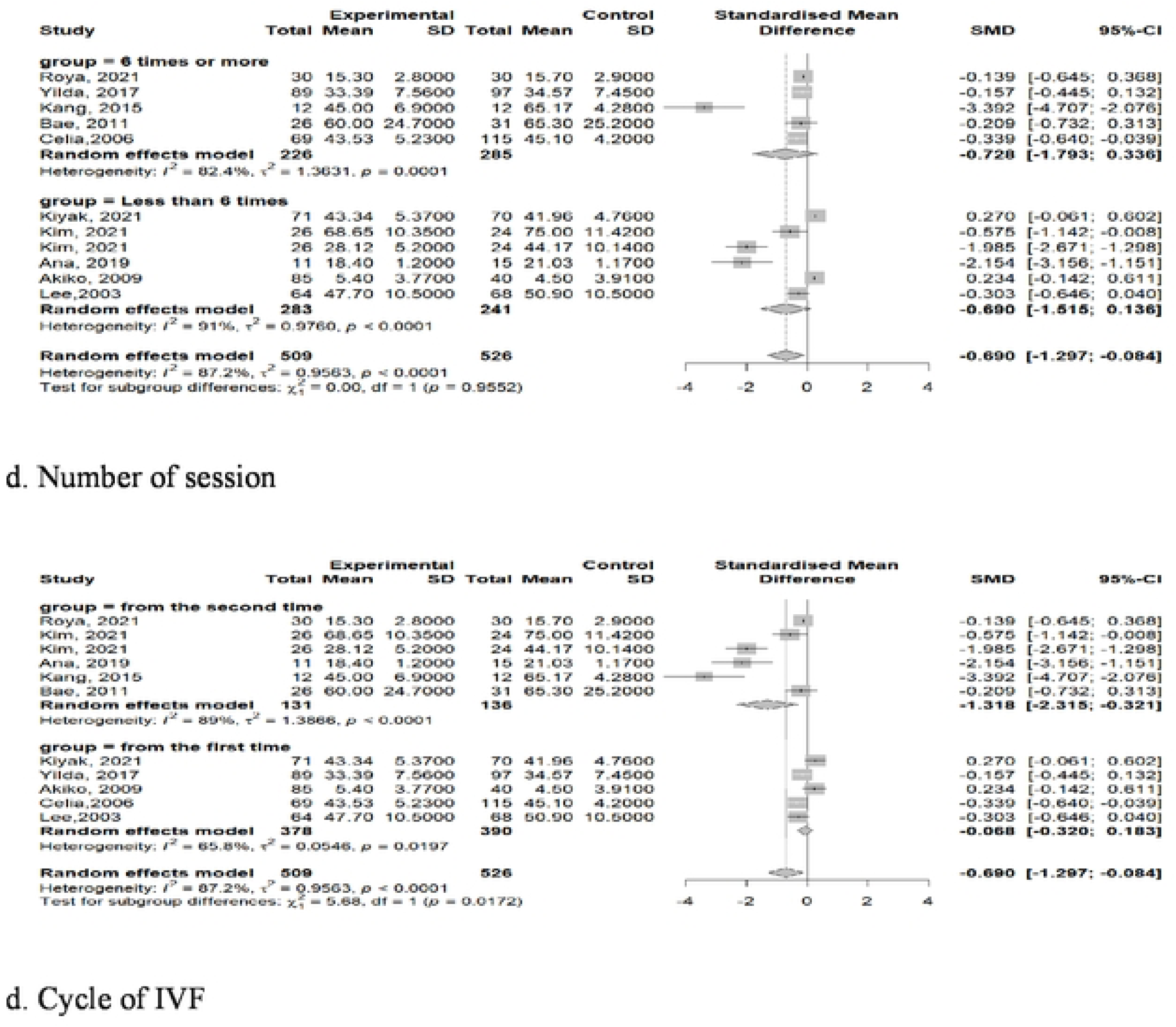
Forest plots for analysis of the moderator effect of relaxation-intervention program.

**Figure 5.**
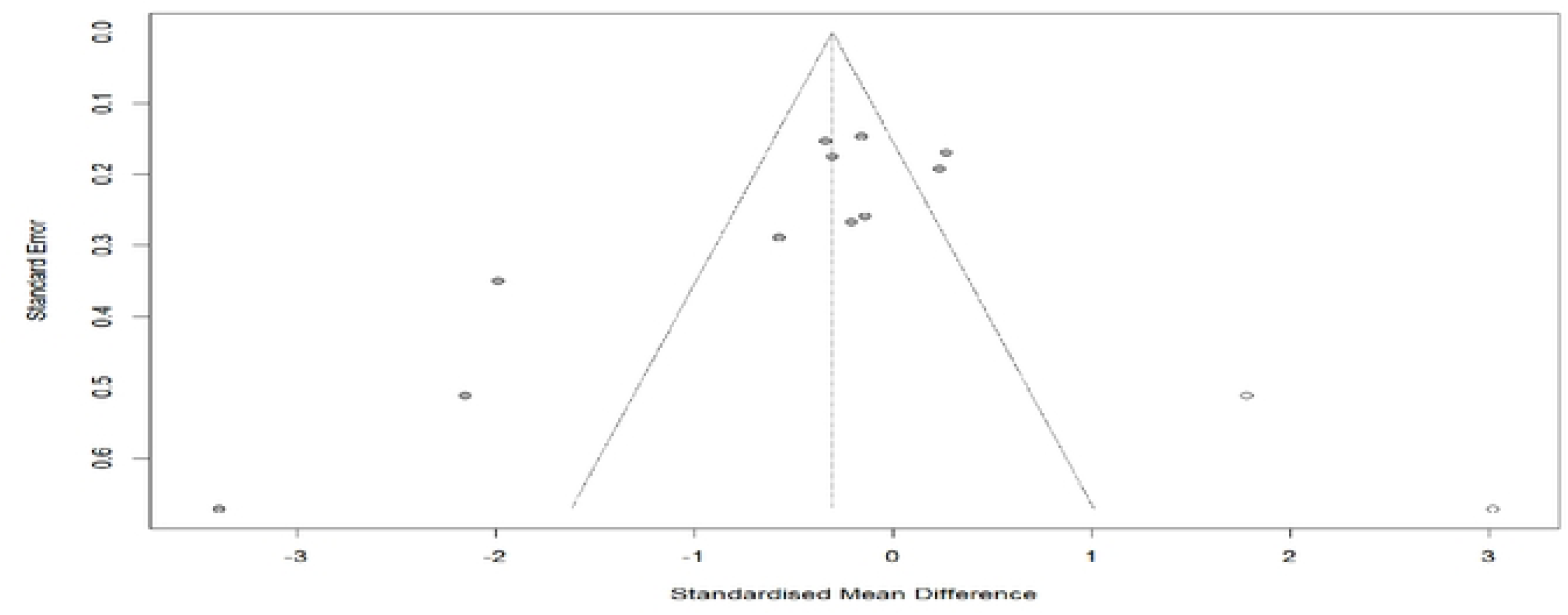
Result of publication bias analysis.

When study design was examined as a moderator, the pooled effect size for RCTs was Hedges’ *g* = −0.63 (95% CI: −1.42 to 0.16), which was statistically non-significant, and substantial heterogeneity was observed (*I*² = 86.3%; *p* < .0001). For NRCTs, the pooled effect size was Hedges’ *g* = −0.90 (95% CI: −1.95 to 0.14), which was also non-significant, with similarly high heterogeneity (*I*² = 88.2%; *p* = .001). The test for subgroup differences indicated no statistically significant difference in effect sizes between RCTs and NRCTs (*Q*b = 0.17; *df* = 1; *p* = .682).

When delivery mode was analyzed as a moderator, group-based interventions showed a pooled effect size of Hedges’ *g* = −0.24 (95% CI: −1.70 to 1.22), which was non-significant, with no observed heterogeneity (*I*² = 0.0%). In contrast, individually delivered interventions demonstrated a statistically significant reduction in anxiety, with a pooled effect size of Hedges’ *g* = −0.81 (95% CI: −1.51 to −0.10), although heterogeneity remained substantial (*I*² = 89.7%). However, the test for subgroup differences indicated that the difference in effect sizes between group-based and individual interventions was statistically non-significant (*Q*b = 0.47; *df* = 1; *p* = .494).

When intervention duration was examined as a moderator, interventions lasting 40 min or less showed a small, non-significant effect on anxiety (Hedges’ *g* = −0.17; 95% CI: −0.42 to 0.08). In this subgroup, heterogeneity was low (*I*² = 0.0%). Similarly, the subgroup with intervention durations of > 40 min to 100 min showed a pooled effect size of Hedges’ *g* = −0.50 (95% CI: −1.45 to 0.45), which was non-significant, and heterogeneity remained high (*I*² = 91.5%). In contrast, for interventions lasting > 100 min, the pooled effect size was Hedges’ *g* = −1.48 (95% CI: −2.78 to −0.16), which was statistically significant, with high heterogeneity (*I*² = 91.5%). Overall, differences in effect size according to intervention duration were statistically non-significant (*Q*b = 3.99; *df* = 2; *p* = .136).

When the number of intervention sessions was examined as a moderator, interventions delivered in six sessions or more showed a pooled effect size of Hedges’ *g* = −0.73 (95% CI: −1.79 to 0.34), which was non-significant, with high heterogeneity (*I*² = 82.4%). In contrast, interventions delivered in fewer than six sessions demonstrated a pooled effect size of Hedges’ *g* = 0.69 (95% CI: −1.52 to 0.14), which was also non-significant, and heterogeneity remained substantial (*I*² = 91.0%). The difference in effect sizes between the two subgroups was statistically non-significant (*Q*b = 0.00; *df* = 1; *p* = .96).

When the number of IVF cycles was examined as a moderator, women undergoing a second or subsequent IVF cycle showed a substantial and statistically significant reduction in anxiety (Hedges’ *g* = −1.32; 95% CI: −2.32 to −0.32). Heterogeneity in this subgroup was high (*I*² = 89.0%). In contrast, women undergoing their first IVF cycle demonstrated a negligible and non-significant effect on anxiety (Hedges’ *g* = −0.07; 95% CI: −0.33 to 0.18), with moderate heterogeneity (*I*² = 65.8%). The difference in effect sizes between the two subgroups was statistically significant (*Q*b = 5.68; *df* = 1; *p* = .017), indicating that the effectiveness of mind–body relaxation interventions differed according to the number of prior IVF cycles.

### 3.5. Publication Bias Analysis

Publication bias was assessed using funnel plot analysis for anxiety outcomes only, as recommended by Higgins and Green [31] when the number of effect sizes is 10 or more. Visual inspection suggested funnel plot asymmetry; therefore, the trim-and-fill method was applied. Two potentially missing effect sizes were imputed, yielding an adjusted pooled effect size of Hedges’ g = −0.30 (95% CI: −1.11 to 0.50), which was comparable to the observed pooled effect size (Hedges’ g = −0.47). The magnitude and direction of the effect estimate remained essentially unchanged after adjustment, indicating that publication bias had a limited impact on the overall findings. Substantial heterogeneity was observed across studies (I² = 89.7%; Q = 116.80; df = 12; p < .01). Although both Egger’s linear regression test (t = −3.74; p < .01) and Begg’s rank correlation test (z = −2.41; p = .01) indicated potential publication bias, sensitivity analysis confirmed that no single study disproportionately influenced the pooled effect size, supporting the robustness of the results.

## 4. Discussion

This systematic review and meta-analysis evaluated the effectiveness of mind–body relaxation programs for women with infertility undergoing assisted reproductive technology (ART). Based on data from 13 studies comprising 1,215 participants, relaxation interventions were associated with significant improvements in anxiety (Hedges’ *g* = −0.69; *p* < .01), depression (Hedges’ *g* = −0.38; *p* < .01), and quality of life (Hedges’ *g* = 0.25; *p* < .01), whereas no statistically significant effect was observed for stress.

These findings align with previous research indicating that psychological distress during infertility treatment is predominantly characterized by anxiety and depression [46, 47]. Specifically, Frederiksen et al. [47] reported small-to-moderate effect sizes for psychosocial interventions targeting patients with infertility, with effect sizes of *d* = 0.31 for anxiety and *d* = 0.38 for depression, supporting the results of the present study.

Hämmerli et al. [48] observed small-to-moderate effects of group psychotherapy on anxiety (*d* = −0.36) and depression (*d* = −0.48), highlighting the potential utility of group-based psychological interventions. In contrast, the present meta-analysis—using Hedges’ *g* and a random-effects model—found that statistically significant reductions in anxiety were observed only in individually delivered interventions, whereas group-based interventions did not demonstrate a significant effect. Although the difference between subgroups was statistically non-significant (*p* = .494), the divergent pattern of effect estimates suggests that the delivery mode may influence psychological outcomes. Given the private and emotionally sensitive nature of infertility, individualized interventions may facilitate greater personalization and psychological safety, thereby addressing participants’ needs more effectively. Consequently, these findings provide differentiated evidence regarding the comparative effectiveness of intervention formats.

Regarding quality of life, Matthiesen et al. [49] reported that while psychological interventions did not significantly improve pregnancy rates, they were effective in enhancing psychological well-being. The small but statistically significant effect size for quality of life identified in the present study (Hedges’ *g* = 0.25) is consistent with these findings, suggesting that relaxation programs contribute to overall improvements in quality of life beyond symptom reduction. However, as noted by Verhaak et al. [50], heterogeneity across studies may be attributable to variations in measurement instruments, particularly the use of fertility-specific tools (e.g., FertiQoL) versus generic measures (e.g., SF-36).

The number of *in vitro* fertilization (IVF) cycles was identified as a statistically significant moderator (*p* = .017). Women undergoing a second or subsequent IVF cycle demonstrated a substantial and statistically significant intervention effect (Hedges’ *g* = −1.32), whereas women undergoing their first IVF cycle showed a negligible and non-significant effect (Hedges’ *g* = −0.07). These findings indicate that women with a history of repeated IVF cycles derive greater psychological benefit from relaxation programs than those undergoing IVF for the first time. This pattern is consistent with evidence suggesting that psychological distress compounds with repeated treatment failure. Peterson et al. [51] reported that depressive symptoms and infertility-related stress increase with the number of failed IVF cycles, with women experiencing two or more failures exhibiting a 3.7-fold higher risk of clinically significant depression. Similarly, Pasch et al. [52] found that psychological distress did not significantly influence treatment outcomes among women undergoing their first IVF cycle, which aligns with the minimal intervention effect observed in first-cycle participants in the present study. In contrast, among women with repeated treatment failure, psychological interventions may contribute to symptom reduction, improved treatment adherence, and sustained engagement with subsequent cycles [53]. These findings suggest that psychological interventions should not be applied uniformly; rather, targeted and intensive relaxation programs should be prioritized for high-risk subgroups, particularly women with a history of repeated IVF failure. In the moderator analysis based on session duration, the pooled effect size increased with longer intervention sessions. Interventions lasting > 100 min showed the largest effect (Hedges’ *g* = −1.48), followed by those lasting 40 to 100 min (Hedges’ *g* = −0.50), whereas sessions shorter than 40 min demonstrated a small, non-significant effect (Hedges’ *g* = −0.17). Although the difference in effect sizes across subgroups was statistically non-significant (*p* = .136), heterogeneity was minimal in the < 40 min subgroup (*I*² = 0.0%), suggesting that brief interventions—such as music therapy or simple breathing exercises—were relatively standardized across studies. In contrast, substantial heterogeneity was observed in sessions exceeding 100 min (*I*² = 91.5%), reflecting variability in intervention content, including mindfulness-based meditation, cognitive behavioral components, and body-oriented practices such as yoga. Clinically, while brief sessions may induce short-term relaxation, longer sessions may provide greater opportunities for psychological processing and skill acquisition. This pattern is consistent with established frameworks such as the Mindfulness-Based Stress Reduction (MBSR) program, which recommends session durations of approximately 2.5 h to allow adequate time for practice and integration [54].

Regarding the number of intervention sessions, neither subgroup showed a statistically significant effect on anxiety. Interventions delivered in six or more sessions yielded a pooled effect size of Hedges’ *g* = −0.73, while those delivered in fewer than six sessions showed a pooled effect size of Hedges’ *g* = −0.69. The difference between subgroups was statistically non-significant (*p* = .96), indicating that the number of sessions did not significantly moderate the effect of relaxation programs. This implies that intervention effectiveness may depend less on the quantity of sessions and more on qualitative factors such as intervention fidelity, facilitator expertise, and participant engagement. Additionally, the timing of outcome assessment—typically conducted immediately post-intervention—may have limited the detection of cumulative effects in longer programs.

In the analysis by study design, both randomized controlled trials (RCTs) and non-randomized controlled trials (NRCTs) demonstrated reductions in anxiety; however, neither subgroup showed a statistically significant pooled effect. Although NRCTs yielded a larger effect size magnitude than RCTs (Hedges’ *g* = −0.90 vs. −0.63), this finding requires cautious interpretation. Intervention effects may be overestimated in non-randomized designs due to selection bias [38]. In NRCTs, participants with higher baseline motivation may be more likely to adhere to the intervention, potentially inflating observed effects. Given that the overall conclusions of this meta-analysis are largely informed by RCT evidence, the findings are considered methodologically robust despite the observed heterogeneity.

Overall, this meta-analysis demonstrates that mind–body relaxation interventions confer psychological benefits for women undergoing ART, particularly regarding anxiety and depressive symptoms. Although heterogeneity across studies was substantial, consistent patterns emerged in moderator analyses. Greater effects were observed among women with repeated IVF failure and in individually delivered interventions, suggesting that psychological vulnerability and delivery format influence effectiveness. These findings indicate that psychological support in infertility care is most beneficial when selectively targeted rather than implemented universally. Tailored, individualized interventions appear particularly appropriate for women experiencing sustained treatment-related distress. Future research should prioritize methodologically rigorous RCTs with standardized outcome measures and longer follow-up periods to evaluate the sustainability of intervention effects and their potential influence on reproductive outcomes.

## 5. Conclusion

This meta-analysis indicates that mind–body relaxation interventions are associated with improvements in anxiety, depression, and quality of life among women undergoing ART, with more pronounced benefits observed among women with repeated IVF failure and when interventions are delivered individually. These findings underscore the importance of targeted, rather than universal, psychological support during fertility treatment. Despite heterogeneity across studies, evidence from randomized trials supports the clinical utility of mind–body relaxation as an adjunct to fertility care. Integrating tailored, individualized relaxation programmes into routine fertility services may enhance psychological well-being and support woman-centered infertility care.

## Data Availability

All relevant data are within the manuscript and its Supporting Information files.

## [Declarations]

### Author Contributions

Conceptualization: SA, Park

Methodology: SA, Park, HY, Kim

Literature search and screening: SA, Park, HY, Kim

Data extraction: SA, Park

Formal analysis: SA, Park

Writing – original draft: SA, Park

Writing – review & editing: SA, Park, HY, Kim

Supervision: HY, Kim

### Ethical statement

Ethical approval was not required for this systematic review.

## Acknowledgments

None

## Declaration of AI use

AI not used in the preparation of this work

